# Liver Volumetry from Magnetic Resonance Images with Convolutional Neural Networks

**DOI:** 10.1101/2021.12.01.21267152

**Authors:** Sara L. Saunders, Justin M. Clark, Kyle Rudser, Anil Chauhan, Justin R. Ryder, Patrick J. Bolan

## Abstract

Measurements of liver volume from MR images can be valuable for both clinical and research applications. Automated methods using convolutional neural networks have been used successfully for this using a variety of different MR image types as input. In this work, we sought to determine which types of magnetic resonance images give the best performance when used to train convolutional neural networks for liver segmentation and volumetry.

Abdominal MRI scans were performed at 3 Tesla on 42 adolescents with obesity. Scans included Dixon imaging (giving water, fat, and T2* images) and low-resolution T2-weighted anatomical scans. Multiple convolutional neural network models using a 3D U-Net architecture were trained with different input images. Whole-liver manual segmentations were used for reference. Segmentation performance was measured using the Dice similarity coefficient (DSC) and 95% Hausdorff distance. Liver volume accuracy was evaluated using bias, precision, and normalized root mean square error (NRMSE).

The models trained using both water and fat images performed best, giving DSC = 0.94 and NRMSE = 4.2%. Models trained without the water image as input all performed worse, including in participants with elevated liver fat. Models using the T2-weighted anatomical images underperformed the Dixon-based models, but provided acceptable performance (DSC ≥ 0.92, NMRSE ≤ 6.6%) for use in longitudinal pediatric obesity interventions. The model using Dixon water and fat images as input gave the best performance, with results comparable to inter-reader variability and state-of-the-art methods.

## 1. Introduction

Making accurate measurements of liver volume is valuable for several clinical and research needs. Preparation for bariatric surgery often involves prescribed weight loss intended to reduce liver volume, which can simplify the surgical procedure and improve some outcomes (1–3). Accurate volumetry is also essential for assessing potential living liver donors to ensure that an appropriate graft size can be collected while leaving a sufficient liver remnant (4). Research studies focusing on non-alcoholic fatty liver disease (NAFLD) can benefit from quantitative measurements of liver size and fat content. NAFLD is the most prevalent chronic liver disease, affecting ∼25% of adults worldwide (5). Early stages of NAFLD may be asymptomatic, but later stages may progress to liver failure (6). NAFLD is also associated with obesity, and in adults weight loss has been associated with a decrease in both liver volume and fat content (7–9). Volume measurements are a valuable adjunct to fat content measurements because, while related, they are independent measures of the liver condition and its response to treatment (10).

Manual delineation of liver borders on either CT or MR images is currently the standard method for determining liver volume, even though it is time consuming and subjective. Methods to automate liver segmentation have long been a subject of research. Past competitions such as SLIVER07 (11) encouraged participants to develop algorithms to accurately segment the liver from publicly available CT images, with the winning entrant giving volume estimation error of −2.9% and a Dice similarity coefficient (DSC, a measure of volume overlap) of 0.97. Segmentation based on MR images has generally been a more difficult problem. Methods have been developed to work with a variety of MR image types as input, including anatomical images (12,13), quantitative T1 or T2* maps (14,15), and water images reconstructed from multi-echo Dixon imaging (also called chemical shift encoded imaging) (16). Techniques using convolutional neural networks (CNNs) based on the U-Net architecture (17,18) have generally shown the best performance. The *no-new U-Net* technique (19) won the liver MR volume task of the recent CHAOS challenge (12) with a DSC of 0.95 and error of 2.85%, using either T_1_-weighted anatomical images or Dixon in/out of phase images as input. In an analysis of the Hepat1ca dataset (20), which included quantitative T1 and T2* maps, Owler et al. used a 3D U-Net to produce a median DSC of 0.97 (15).

Our long-term goal is to use liver volumetry in longitudinal obesity treatment studies among adolescents. Studies in adults have shown changes in liver fat content and volume with dietary, medical, and surgical interventions (7,21,22). Such studies commonly use Dixon imaging to quantify liver steatosis (23). Dixon imaging methods generate multiple image contrasts from a single acquisition, including water, fat, T2* or R2* maps, and maps of the proton density fat fraction (PDFF). Based on prior work it is evident that these can be used as input for CNNs to segment the liver and estimate its volume, but it is not clear which image or combination of images would give the best performance. A water image alone can be used, and can give volumes that are not greatly biased by the degree of fat content (16). However, these images have low contrast where the liver adjoins the chest wall, stomach, and heart, and therefore it may be possible to improve performance by adding additional contrasts.

The primary goal of this work is to compare the performance of CNNs trained with different images derived from Dixon acquisitions to determine the best-performing set of input images. A secondary goal was to determine if the anatomical images acquired during these studies could be used to train CNNs with comparable performance. Low-resolution T2-weighted images are commonly acquired for scout imaging in both clinical and research settings, but these have highly anisotropic spatial resolution and often feature blurring and banding artifacts. If these image series could provide accurate volume estimates it would enable analyses of large retrospective image series that were not prospectively designed to include quantitative Dixon scans. In this study, we trained CNNs using both Dixon and anatomical images as inputs, and compared their performance using standard segmentation metrics, liver volume accuracy, and evaluated their sensitivity to liver fat content. This manuscript extends the initial findings we reported in an earlier conference abstract (24).

## 2. Material and Methods

A total of 42 adolescents (age 13-18 years) with obesity (body mass index > 95^th^ percentile by age and sex) were consented and completed a clinical trial approved by our Institutional Review Board (clinicaltrials.gov NCT02496611). Participants were imaged prior to any intervention on a 3 T Siemens Prisma^*fit*^ MR system (Siemens, Erlangen, Germany) using standard spine and body receive coils. Participants received repeated scans in the study, but only the baseline MRI scan for each subject was used for this analysis.

Imaging included two single breath-hold T2-weighted scans used for anatomical reference: an axial balanced steady-state free precession (TrueFISP) scan (TR/echo time = 399/1.56 ms, flip angle 60°, matrix 320×260, FOV 440×360 mm, 8.0 mm slice thickness, 40 slices with a 20% gap, GRAPPA acceleration R=2, readout bandwidth 1420 Hz/pixel, reconstructed to 1.4 × 1.4 mm in-plane, 18 s acquisition time) and a coronal half-Fourier acquisition single-shot turbo spin-echo (HASTE) scan (TR/echo time = 1000/86 ms, flip angles 90/124°, matrix 256×256, FOV 440×440 mm, 8.0 mm slice thickness, 17 slices with a 75% gap, GRAPPA acceleration R=3, readout bandwidth 698 Hz/pixel, reconstructed to 1.7 × 1.7 mm in-plane, 17 s acquisition time). Liver fat was quantified using a single breath-hold 3D Dixon scan with six echo times (TE=1.15-7.23 ms, ΔTE=1.23 ms, TR=9ms, flip angle = 4°, matrix 224×198×64, FOV 450×398×224 mm, CAIPIRINHA acceleration R=2×2, readout bandwidth 1060 Hz/pixel, reconstructed to 2 × 2 × 3.5 mm, 18 s acquisition time). From this acquisition the water (W), fat (F), fat fraction (FF), and T2-star (T2*) images were reconstructed using the vendor’s *qdixon* software.

To provide objective measurements of liver volume, the Dixon source images were deidentified and sent to a commercial provider of medical imaging processing services (3DR Labs, LLC, Louisville KY) with expertise in liver segmentation. For all datasets the liver boundaries were manual delineated on each axial slice containing liver tissue. These manual whole-liver segmentations were used as a ground truth for calculating liver volumes and fat fraction (by averaging FF over the volume) and for training CNNs.

The coronal HASTE and axial TrueFISP images required interpolation and registration to align them with the Dixon images and manual segmentations. Preprocessing included interpolation of the images to match the Dixon resolution, manual cropping to reduce regions beyond the liver, and coarse manual shifting in the superior-inferior direction to correct for inconsistent breath holds. Registration was performed with an affine registration followed by a multi-resolution non-rigid B-spline registration, implemented in Python using the SimpleElastix (25) library. The registration used a gradient descent optimizer, mutual information as the agreement metric, and a bending energy penalty to constrain the degree of warping. After reviewing the results of automatic segmentation, five cases required additional warping using 20-40 manually selected control points to produce acceptable registration. Figure 1 shows a set of all images used for this study, including the co-registered anatomical images.

**Figure 1.**
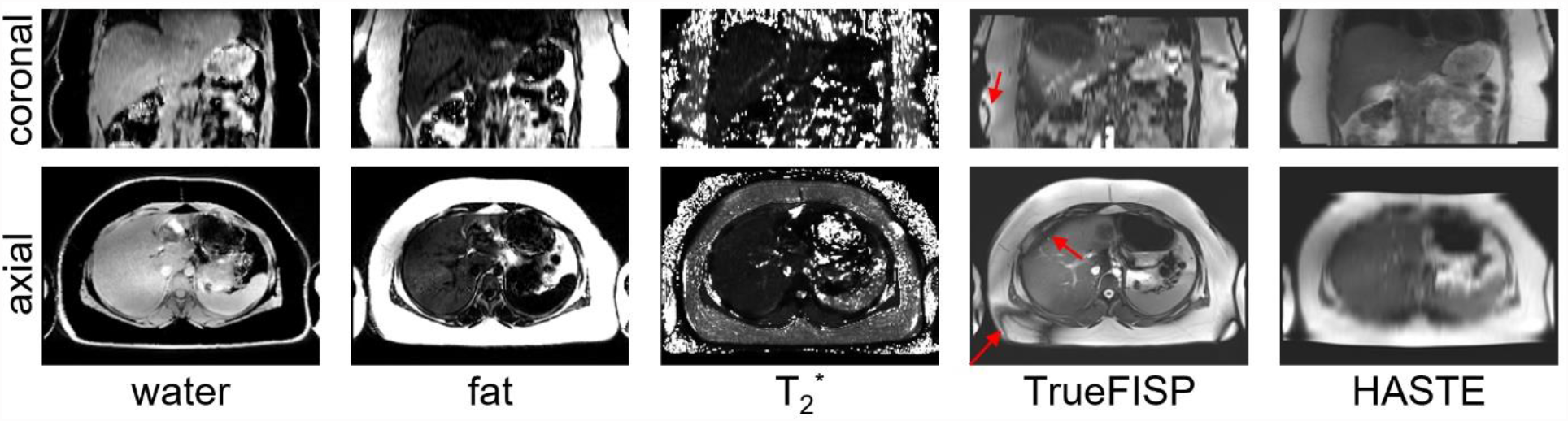
Source images from a subject with NAFLD (FF = 17.7%), reformatted in axial and coronal planes. The TrueFISP and HASTE images are shown after interpolation and registration to align them with the 3D Dixon geometry. Note the low resolution of the TrueFISP in the coronal plane, and HASTE in the axial plane, due to their large slice thickness (8 mm). The TrueFISP images also show the characteristic dark bands due to off-resonance, which are present even in the liver (arrows). Note also low contrast between water and both the chest wall and the heart, a factor which motivated this study.

CNNs were trained to produce liver segmentations using the manual segmentations as ground truth. Seven distinct CNNs were trained using the same ground truth but with different input images. Three networks used a single image type from the Dixon acquisition (W, F, and T2*), two used combinations (WF = water and fat, WFT2* = water, fat, and T2*), which were input as multiple channels. Additionally, two networks were trained using the anatomical images (TrueFISP and HASTE). Note that that PDFF images were not evaluated as input because they can be algebraically derived from W and F, and could result in a redundant network. For brevity, each distinct CNN will be identified by their input image set; e.g., the CNN trained using water and fat as input images will be referred to as the *WF model*.

All modeling, training, and inference was performed in Python using Keras with Tensorflow (26,27). All models used the same 3D U-Net architecture (18), but with varying numbers (1-3) of input channels, as detailed in Figure 2. Training was performed with five-fold cross validation with random augmentation (3D rotation, translation, and scaling) of the input data. Each network was trained over 350 epochs with batch size 2, using DSC as the loss function and Adam (28) as the optimizer with a learning rate of 10^−4^.

**Figure 2.**
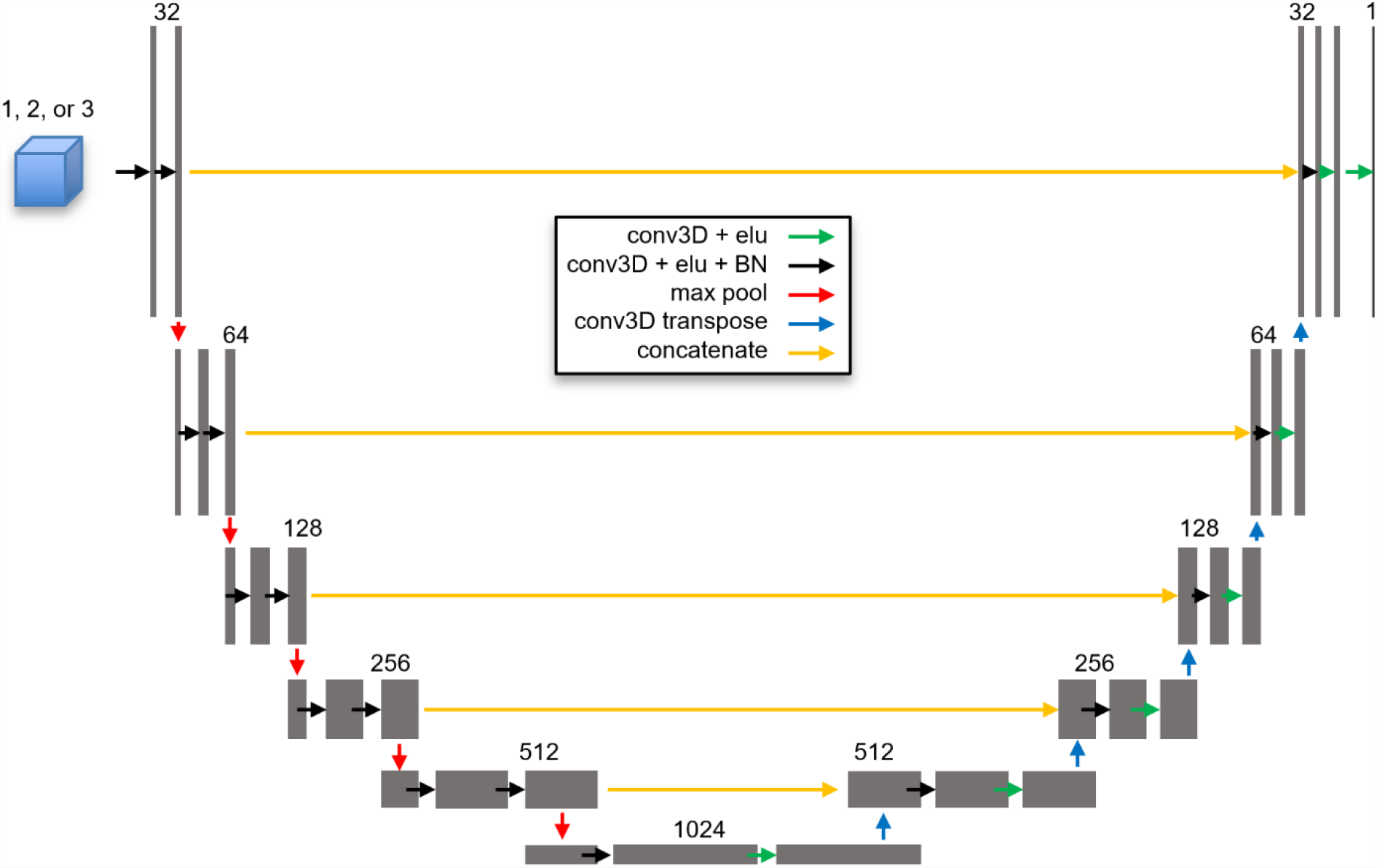
Architecture of the six-layer 3D U-Net used for all models. All models had input image size of 224×224×64, with 1-3 channels depending on the specific model. The number of channels doubled with each layer, varying from 32 to 1024 at the bottom layer.

The performance of the segmentations from each set of image inputs was assessed using DSC, which measures the relative intersection between true and predicted segmentations, and the 95% Hausdorff distance (HD95), which characterizes the distance between true and predicted segmentation boundaries. The accuracy of estimating total liver volume was assessed using both bias (mean error) and precision (standard deviation of error), and also by using the normalized root-mean-square error (NRMSE, RMSE divided by mean ground truth volume) as an aggregate metric of accuracy (29).

Analyses were performed with all data and within mean liver fat content subgroups of normal (mean FF<5.5%) and NAFLD (>= 5.5%). The performance metrics (DSC and HD95) were compared between the outputs of each model, and in all three liver fat content subgroups, using paired t-tests (2 metrics x 3 subgroups x 21 combinations = 126 comparisons in total). Additionally, the performance between NAFLD and normal groups were compared for each metric and model using independent t-tests. Correlations between volumes, body mass index (BMI), and fat fraction were assessed using Pearson’s correlation. All statistical tests were performed in python using scipy 1.6.0. (30). A P value < 0.05 was considered statistically significant.

## 3. Results

In this population of 42 participants, fat fraction ranged from 2.6% to 23.4% (mean 7.9%, standard deviation 5.7%), and BMI ranged from 31.3 to 50.8 kg/m^2^ (mean 39.2, standard deviation 4.9). Twenty-three participants had liver fat <5.5% and were classified as normal, while 19 had ≥ 5.5% and were classified as having NAFLD. Liver volumes based on the manual segmentations ranged from 1327 to 3252 mL (mean 1970.1, standard deviation 421.6 mL), and were moderately correlated with both BMI (R=0.66, p<0.001) and mean fat fraction (R=0.66, p<0.001).

The segmentation performance of all seven models is shown in Figure 3. Considering all subjects, models based on Dixon WF and WFT2* gave the highest DSC values, which were each significantly higher than any other model but not significantly different from each other. These two models also gave significantly lower HD95 values than other models, with the exception that the difference between WF and W did not reach statistical significance (p=0.122). The performance of WF and WFT2* was closely followed by that of W. Figure 3a and 3b show a trend that segmentation performance measured by both DSC and HD95 was a little better in NAFLD cases than in normal liver, although this did not reach statistical significance in all but one comparison (HD95 for the F model, p=0.032). The models based on anatomical images (TrueFISP and HASTE) gave poorer segmentation performance compared to Dixon image models, as expected due to the lower spatial resolution.

**Figure 3.**
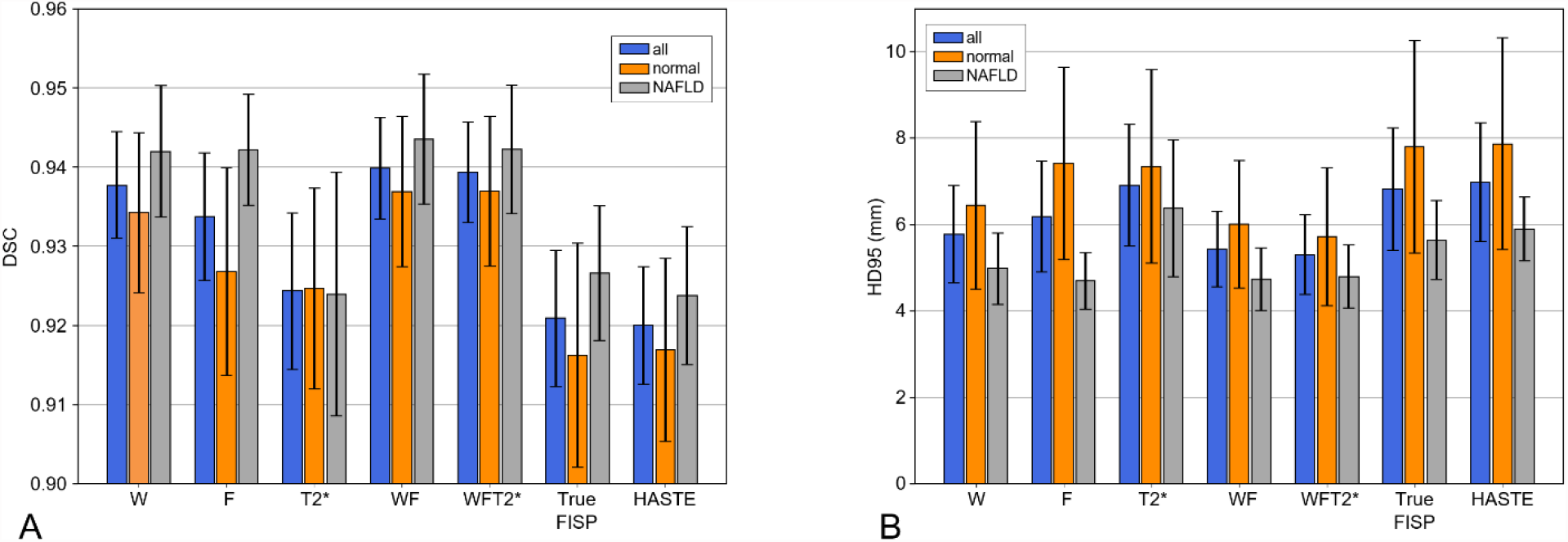
Segmentation performance of CNNs trained with different input images. For both A) Sorensen-Dice scores and B) 95% Hausdorff distance metrics, results are shown for all cases, and subdivided into normal liver and NAFLD (liver fat >= 5.5%). Error bars indicate 95% confidence intervals.

The primary goal of this study was to evaluate accuracy of liver volume measurements, as shown in Figure 4. The NRMSE (Figure 4a) and precision (Figure 4b) results show that the top three models all include the Dixon-W image (W, WF, WFT2*), with no statistically significant differences between them in pairwise t-tests. Note that each of the predicted liver segmentations tend to overestimate the liver volume (Figure 4c), although this effect is small, on the order of 1%. In these accuracy measurements, several of the models have notably different performance depending on the NAFLD status. This is an undesirable characteristic for a predictor of liver volume, as it could introduce bias in a longitudinal study where liver fat levels change over time.

**Figure 4.**
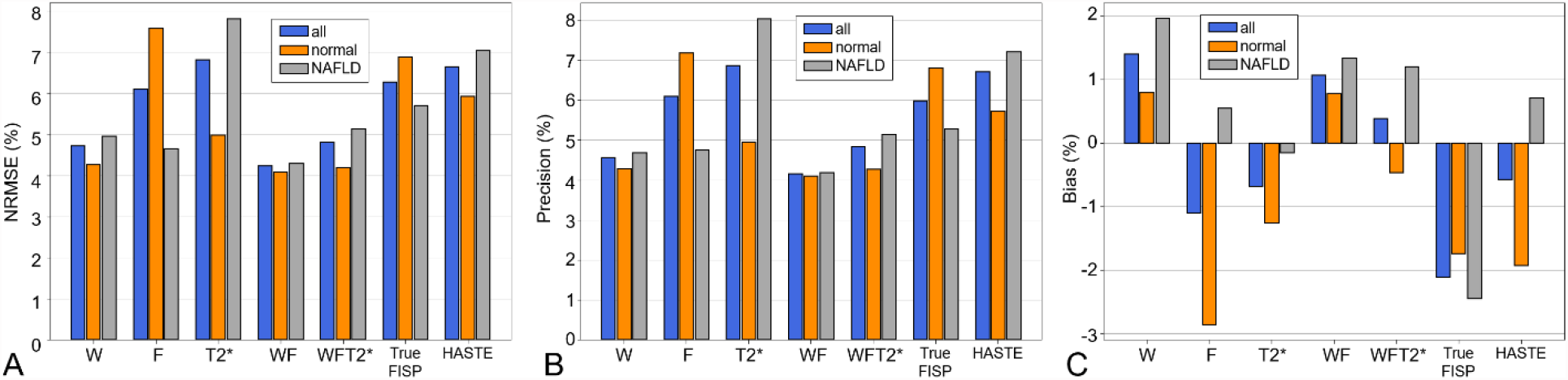
Accuracy of liver volume estimation using networks trained with different input images. A) overall accuracy, represented by the root-mean-square error, B) precision, and C) bias. For each, results are shown for all cases and subdivided into normal liver and NAFLD groups.

Based on segmentation performance and accuracy metrics, it can be argued that the WF model is the most appropriate for estimating liver volumes in longitudinal studies of steatosis, giving overall performance of DSC=0.940, HD95=5.44 mm, NRMSE=4.2%, bias=1.1%, precision=4.1%. This model has slightly higher bias than the WFT2* model (DSC=0.939, HD95=5.31 mm, NRMSE=4.8%, bias=0.4%, precision=4.8%), but it gives greater consistency between normal and NAFLD cases in accuracy metrics.

For the models based on anatomical images, the TrueFISP model (DSC=0.921, HD95=6.82 mm, NRMSE=6.3%, bias=-2.1%, precision=6.0%) generally outperforms the HASTE model (DSC=0.920, HD95=6.98 mm, NRMSE=6.6%, bias=-0.6%, precision=6.7%). While the TrueFISP model underestimates liver volumes by ∼2%, it has slightly better precision and overall accuracy, and its bias does not vary greatly with NAFLD status.

The best-performing models based on segmentation performance (W, WF, and WFT2*) also gave the best performance based on accuracy and precision. However, the overall correlation between segmentation performance and accuracy was modest. Pooling across all models and subjects, the correlation coefficient between NRMSE and DSC was R=0.19 (p=0.001), and R=−0.27 for HD95 (p<0.001).

Two representative examples of segmentation performance with the Dixon-based images are shown in Figure 5. These examples compare the results of the best- and worst-performing models (WF, and T2* respectively) with the gold-standard manual segmentation. The two cases are representative of cases with high (left column, A, B) and low (right column, C, D) accuracy, based on the error across all models. Note that the errors in these cases are predominantly at the borders with the heart and chest wall; this trend was observed throughout the full dataset.

**Figure 5.**
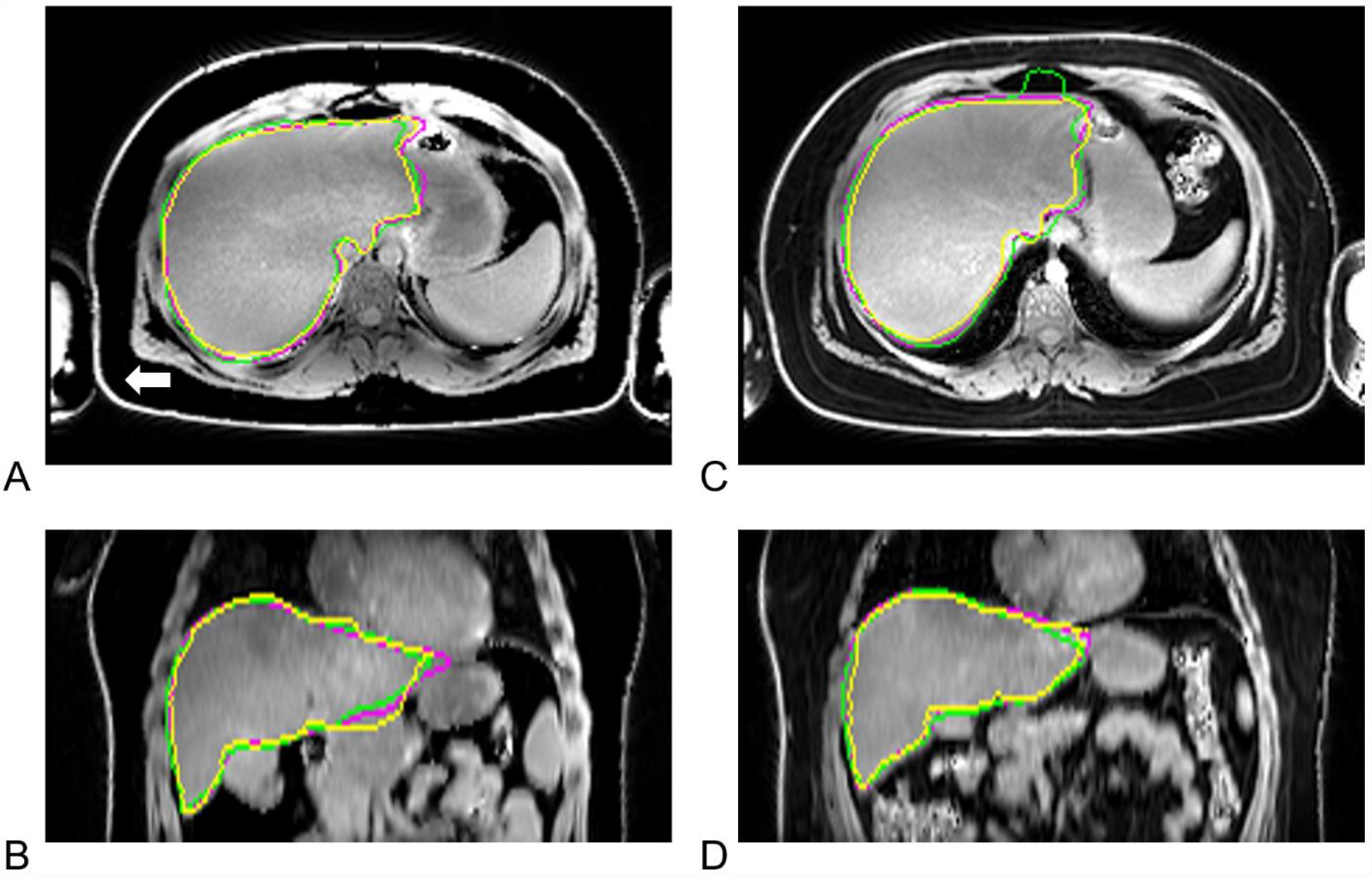
Two examples of segmentation using Dixon-based images. These are axial (A, C) and coronal (B, D) views of a Dixon-W image, overlayed with the manual (ground-truth) segmentation (yellow), the segmentation from the WF (magenta) and T2* (green) models. The examples selected represent the range of accuracy, with the case case on the left (A,B) having high accuracy (WF error 1.6%, DSC=0.96) and the case on the right (C, D) with relatively low accuracy (WF error 10.9%, DSC=0.941).

Figure 6 shows examples of segmentations with the T2-weighted anatomical images. While these models do not perform as well as the Dixon-based models, the segmentations have good overall performance and accuracy despite the low spatial resolution and clear presence of imaging artifacts.

**Figure 6.**
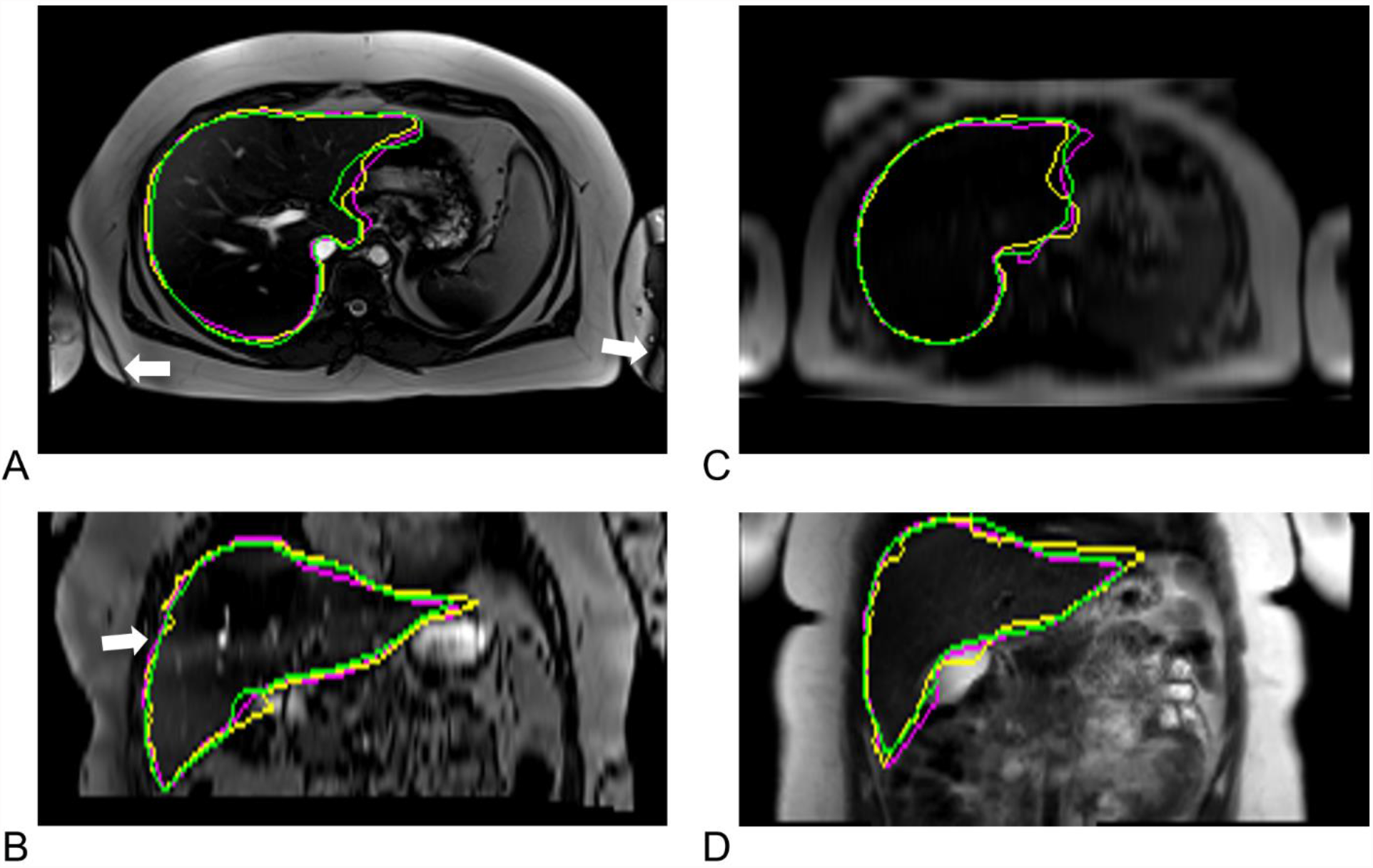
Representative examples of segmentation using anatomical images. Left column shows A) axial and B) coronally reformatted TrueFISP images showing manual (yellow), HASTE (green, DSC=0.935), and TrueFISP (magenta, DSC=0.924) liver boundaries in a subject with 11.9% liver fat, overlayed on the TrueFISP image. Banding from off-resonance can be seen in adipose tissue and in the liver (white arrows). Right column (C, D) shows segmentations overlayed on the HASTE image in subject with 4.0% liver fat, and DSC scores of 0.913 for HASTE and 0.914 for TrueFISP.

## 4. Discussion

The best performing CNNs based on both accuracy and segmentation metrics were the three that included the Dixon-W image as input: W, WF, and WFT2*. Relative to using just the W image, adding F and T2* images to the input set gave modest improvement in segmentation performance but no statistically significant improvement in accuracy. Using multiple Dixon images as model inputs adds a minor complexity and slightly increases training duration, but does provide a modest performance benefit. Of all the models evaluated, the WF model would be most useful for longitudinal studies which may have changes in liver fat content over time, as this model had the smallest bias difference between subjects with and without NAFLD. These models had moderate but non-statistically significant performance differences for normal participants and those with NAFLD, similar to the findings reported by Stoker et al. which used Dixon-W as input for their model (16).

The overall performance of the WF model in this work (DSC=0.94, error 4.2%) was comparable but slightly lower than that of state-of-the-art nnUNet (DSC=0.95, error 2.9%), and that of human inter-reader performance (DSC=0.95, error 1.5%), as reported in the CHAOS challenge data set (12). This can be partly attributed to the smaller training dataset used in this study (n=42 vs n=60). This level of performance is suitable for use in longitudinal studies of NAFLD treatments. Using manually segmentation, Luo et al. reported liver volume decreases of 12.2% over a 2-week liquid diet, and 21% from baseline to 1-month post bariatric surgery (7). Notably, in the 6 months post-surgery, liver volume was unchanged while PDFF continued to decrease, underscoring the independence of PDFF and volume measurements.

While the performance of models based on T2-weighted anatomical images was lower than that of the Dixon-based models, the segmentation performance (DSC≥0.92) and volume accuracy (NMRSE≤6.6%) were better than expected. Despite the thick slices, gaps between slices, and substantial blurring (in HASTE) and banding artifacts (in TrueFISP), the trained models were still able to produce good volumetric segmentations. These results can be used to design retrospective studies of larger clinical datasets which do not contain high-resolution anatomical images or Dixon acquisitions.

This study had several limitations. All images were acquired in a pediatric population, and the results may not generalize to an older population with a greater incidence of iron retention conditions, fibrosis, or other liver disease. Furthermore, this comparison used a single CNN architecture for consistent comparisons between input image types. While this architecture is widely used for medical image segmentation, other architectures may lead to different relative performance between the input image types. Finally, the ground truth segmentations used in this work were generated by one reader per case. Our review of these segmentations found them to be of good quality, but this ground-truth could be strengthened by merging segmentations from multiple readers.

## 5. Conclusions

In conclusion, this work compared the liver segmentation performance of CNNs trained with different sets of MR images. A model using Dixon W and F as input gave the best performance (DSC=0.94, error 4.2%), with results comparable to inter-reader variability and state-of-the-art methods.

## Data Availability

All data produced in the present study are available upon reasonable request to the authors

## Abbreviations

BMI: body mass index
CAIPIRINHA: Controlled Aliasing in Parallel Imaging Results in Higher Acceleration
CNN: Convolutional neural network
CT: computed tomography
DSC: Dice similarity coefficient
FOV: Field of view
GRAPPA: GeneRalized Autocalibrating Partial Parallel Acquisition
HASTE: half-Fourier single-shot turbo spin echo
HD95: 95^th^ percentile Hausdorff distance
MRI: magnetic resonance imaging
NAFLD: non alcoholic fatty liver disease
NRMSE: normalized root mean square error
PDFF: proton density fat fraction

## Notes

**Funding:** This work was supported in part by NIH NIDDK R01DK105953, NIH NIBIB P41EB027061, and NIH NCATS UL1TR002494.

### Competing Interest Statement

The authors have declared no competing interest.

### Clinical Trial

NCT02496611

### Funding Statement

This work was supported in part by NIH NIDDK R01DK105953, NIH NIBIB P41EB027061, and NIH NCATS UL1TR002494.

### Author Declarations

IRB of the University of Minnesota gave ethical approval for this work

### Summary of Updates

Reformatted abstract, moved fig S1 into paper, removed note regarding MRM submission, minor layout edits

## References

1. van Wissen J, Bakker N, Doodeman HJ, Jansma EP, Bonjer HJ, Houdijk APJ. Preoperative Methods to Reduce Liver Volume in Bariatric Surgery: a Systematic Review. OBES SURG 2016:6.

2. Romeijn MM, Kolen AM, Holthuijsen DDB, et al. Effectiveness of a Low-Calorie Diet for Liver Volume Reduction Prior to Bariatric Surgery: a Systematic Review. Obes. Surg. 2021;31:350–356 doi: 10.1007/s11695-020-05070-6.

3. Tewksbury C, Crowley N, Parrott JM, et al. Weight Loss Prior to Bariatric Surgery and 30-Day Mortality, Readmission, Reoperation, and Intervention: an MBSAQIP Analysis of 349,016 Cases. Obes. Surg. 2019;29:3622–3628 doi: 10.1007/s11695-019-04041-w.

4. Suzuki K, Epstein ML, Kohlbrenner R, et al. Quantitative Radiology: Automated CT Liver Volumetry Compared With Interactive Volumetry and Manual Volumetry. Am. J. Roentgenol. 2011;197:W706–W712 doi: 10.2214/AJR.10.5958.

5. Younossi ZM, Koenig AB, Abdelatif D, Fazel Y, Henry L, Wymer M. Global epidemiology of nonalcoholic fatty liver disease-Meta-analytic assessment of prevalence, incidence, and outcomes: HEPATOLOGY, Vol. XX, No. X 2016. Hepatology 2016;64:73–84 doi: 10.1002/hep.28431.

6. Byrne CD, Targher G. NAFLD: A multisystem disease. J. Hepatol. 2015;62:S47–S64 doi: 10.1016/j.jhep.2014.12.012.

7. Luo RB, Suzuki T, Hooker JC, et al. How bariatric surgery affects liver volume and fat density in NAFLD patients. Surg. Endosc. 2018;32:1675–1682 doi: 10.1007/s00464-017-5846-9.

8. Edholm D, Kullberg J, Karlsson FA, Haenni A, Ahlström H, Sundbom M. Changes in liver volume and body composition during 4 weeks of low calorie diet before laparoscopic gastric bypass. Surg. Obes. Relat. Dis. Off. J. Am. Soc. Bariatr. Surg. 2015;11:602–606 doi: 10.1016/j.soard.2014.07.018.

9. Holderbaum M, Casagrande DS, Sussenbach S, Buss C. Effects of very low calorie diets on liver size and weight loss in the preoperative period of bariatric surgery: a systematic review. Surg. Obes. Relat. Dis. 2018;14:237–244 doi: 10.1016/j.soard.2017.09.531.

10. Bian H, Hakkarainen A, Zhou Y, Lundbom N, Olkkonen VM, Yki-Järvinen H. Impact of non-alcoholic fatty liver disease on liver volume in humans: Liver volume in humans. Hepatol. Res. 2015;45:210–219 doi: 10.1111/hepr.12338.

11. Heimann T, van Ginneken B, Styner MA, et al. Comparison and Evaluation of Methods for Liver Segmentation From CT Datasets. IEEE Trans. Med. Imaging 2009;28:1251–1265 doi: 10.1109/TMI.2009.2013851.

12. Kavur AE, Gezer NS, Bariş M, et al. CHAOS Challenge - combined (CT-MR) healthy abdominal organ segmentation. Med. Image Anal. 2021;69:101950 doi: 10.1016/j.media.2020.101950.

13. Mulay S, Deepika G, Jeevakala S, Ram K, Sivaprakasam M. Liver Segmentation from Multimodal Images Using HED-Mask R-CNN. In: Li Q, Leahy R, Dong B, Li X, editors. Multiscale Multimodal Medical Imaging. Vol. 11977. Lecture Notes in Computer Science. Cham: Springer International Publishing; 2020. pp. 68–75. doi: 10.1007/978-3-030-37969-8_9.

14. Irving B, Hutton C, Dennis A, et al. Deep Quantitative Liver Segmentation and Vessel Exclusion to Assist in Liver Assessment. In: Valdés Hernández M, González-Castro V, editors. Medical Image Understanding and Analysis. Vol. 723. Communications in Computer and Information Science. Cham: Springer International Publishing; 2017. pp. 663–673. doi: 10.1007/978-3-319-60964-5_58.

15. Owler J, Irving B, Ridgeway G, Wojciechowska M, McGonigle J, Brady SM. Comparison of Multi-atlas Segmentation and U-Net Approaches for Automated 3D Liver Delineation in MRI. In: Zheng Y, Williams BM, Chen K, editors. Medical Image Understanding and Analysis. Communications in Computer and Information Science. Cham: Springer International Publishing; 2020. pp. 478–488. doi: 10.1007/978-3-030-39343-4_41.

16. Stocker D, Bashir MR, Kannengiesser SAR, Reiner CS. Accuracy of Automated Liver Contouring, Fat Fraction, and R2* Measurement on Gradient Multiecho Magnetic Resonance Images: J. Comput. Assist. Tomogr. 2018;42:697–706 doi: 10.1097/RCT.0000000000000759.

17. Ronneberger O, Fischer P, Brox T. U-Net: Convolutional Networks for Biomedical Image Segmentation. In: Navab N, Hornegger J, Wells WM, Frangi AF, editors. Medical Image Computing and Computer-Assisted Intervention – MICCAI 2015. Lecture Notes in Computer Science. Springer International Publishing; 2015. pp. 234–241.

18. Çiçek Ö, Abdulkadir A, Lienkamp SS, Brox T, Ronneberger O. 3D U-Net: Learning Dense Volumetric Segmentation from Sparse Annotation. In: Ourselin S, Joskowicz L, Sabuncu MR, Unal G, Wells W, editors. Medical Image Computing and Computer-Assisted Intervention – MICCAI 2016. Vol. 9901. Cham: Springer International Publishing; 2016. pp. 424–432. doi: 10.1007/978-3-319-46723-8_49.

19. Isensee F, Kickingereder P, Wick W, Bendszus M, Maier-Hein KH. No New-Net. In: Crimi A, Bakas S, Kuijf H, Keyvan F, Reyes M, van Walsum T, editors. Brainlesion: Glioma, Multiple Sclerosis, Stroke and Traumatic Brain Injuries. Vol. 11384. Cham: Springer International Publishing; 2019. pp. 234–244. doi: 10.1007/978-3-030-11726-9_21.

20. Mole DJ, Fallowfield JA, Kendall TJ, et al. Study protocol: HepaT1ca – an observational clinical cohort study to quantify liver health in surgical candidates for liver malignancies. BMC Cancer 2018;18:890 doi: 10.1186/s12885-018-4737-3.

21. Cunha GM, Correa de Mello LL, Hasenstab KA, et al. MRI estimated changes in visceral adipose tissue and liver fat fraction in patients with obesity during a very low-calorie-ketogenic diet compared to a standard low-calorie diet. Clin. Radiol. 2020;75:526–532 doi: 10.1016/j.crad.2020.02.014.

22. Ozaki A, Yoneda M, Kessoku T, et al. Effect of tofogliflozin and pioglitazone on hepatic steatosis in non-alcoholic fatty liver disease patients with type 2 diabetes mellitus: A randomized, open-label pilot study (ToPiND study). Contemp. Clin. Trials Commun. 2020;17:100516 doi: 10.1016/j.conctc.2019.100516.

23. Reeder SB, Hu HH, Sirlin CB. Proton density fat-fraction: A standardized mr-based biomarker of tissue fat concentration. J. Magn. Reson. Imaging JMRI 2012;36:1011–1014 doi: 10.1002/jmri.23741.

24. Saunders SL, Bolan PJ, Ryder JR. Effect of Multi-Parametric MR Images on Accuracy of U-Net Liver Segmentations. In: Proceedings of the ISMRM Workshop on MRI of Obesity & Metabolic Disorders. Singapore; 2019.

25. Marstal K, Berendsen F, Staring M, Klein S. SimpleElastix: A User-Friendly, Multi-lingual Library for Medical Image Registration. In: 2016 IEEE Conference on Computer Vision and Pattern Recognition Workshops (CVPRW).; 2016. pp. 574–582. doi: 10.1109/CVPRW.2016.78.

26. Chollet, Francois. Keras. https://keras.io/. Accessed April 29, 2019.

27. Developers T. TensorFlow. Zenodo; 2021. doi: 10.5281/zenodo.5645375.

28. Kingma DP, Ba J. Adam: A Method for Stochastic Optimization. 14126980 Cs 2017.

29. Obuchowski NA, Reeves AP, Huang EP, et al. Quantitative imaging biomarkers: A review of statistical methods for computer algorithm comparisons. Stat. Methods Med. Res. 2015;24:68–106 doi: 10.1177/0962280214537390.

30. Virtanen P, Gommers R, Oliphant TE, et al. SciPy 1.0: fundamental algorithms for scientific computing in Python. Nat. Methods 2020;17:261–272 doi: 10.1038/s41592-019-0686-2.

